# Cross-sectional study on Intention to be vaccinated against COVID-19 in Benin and Senegal : a structural equation modelling (SEM)

**DOI:** 10.1101/2023.06.29.23292061

**Authors:** Ibrahima Gaye, Valery Ridde, Elías Avahoundjea, Mouhamadou F. Ba, Jean-Paul Dossoua, Amadou I. Diallo, Adama Faye

**Affiliations:** Institute of Health and Development (ISED), Cheikh Anta Diop University, Dakar, Senegal; CEPED, IRD-University of Paris, ERL INSERM SAGESUD, Paris, France; Center for Research in Human Reproduction and Demography (CERRHUD), Cotonou, Benin

**Keywords:** COVID-19, SEM, intention, vaccination, Benin, Senegal

## Abstract

**Introduction:** Vaccination is considered one of the solutions to the COVID-19 pandemic. However, a small proportion of the population was fully vaccinated in Benin (20.9%) and Senegal (7.6%) by December 2022. This study explores the determinants of intent to vaccinate.

**Methods:** This was a cross-sectional, descriptive, and analytical study of 865 Beninese and 607 Senegalese aged 18 years and older. Marginal quota sampling by age, gender and region was adopted. Data collection, using a survey instrument based on the Random Digit Dialing (RDD) method, was conducted from December 24, 2020, to January 16, 2021, in Senegal and from March 29 to May 14, 2021, in Benin. The questionnaire used the Theory of Planned Behavior (TPB) and the Health Belief Model (HBM). The influence of factors was tested using a structural equation model. All analyses were conducted in R.

**Results:** Results show that a good perception of the benefits of vaccination (β_*sen*_ =0.33***; β_*Ben*_=0.12***), a positive attitude (β_*sen*_=0.22***; β_*Ben*_=0.20***), and sensitivity to subjective norms (β_*sen*_=0.19***; β_*Ben*_=0.32***) positively influence the intention to vaccinate. Low trust in health care providers (β_*sen*_=-0.40***; β_*Ben*_=-0.36***) amplifies the perceived risk of vaccination (β_*sen*_=-0.14***; β_*Ben*_=-0.25***), which negatively impacts intention to vaccinate. Perceived vaccine efficacy was affected by perceived risk (β_*sen*_=-0.12***; β_*Ben*_=-0.05***) of the disease and improved by good apprehension of the benefits of vaccination (β_*sen*_=0.60***; β_*Ben*_=0.13***). Aspects related to behavioral control, vaccine information seeking, efficacy, or fairness did not appear as correlates of vaccine intention (P>0.05).

**Conclusion:** Beninese and Senegalese public health authorities could develop additional intervention strategies to improve immunization coverage by considering these influencing factors, the basis of which could be better understood through subsequent qualitative studies.

## INTRODUCTION

The WHO declared the COVID-19 outbreak a global pandemic on March 11, 2020, and invited States to take immediate measures to limit the infection’s progression and ensure compliance with international health regulations (2005) **[1]**. Thus, many countries have decided to prohibit public gatherings, implementing social distancing and especially containment. At the same time, vaccines against COVID-19 are being developed **[2]**. Thus, the WHO has authorized **[3]**, for emergency use, Pfizer (December 31, 2020), AstraZeneca (February 15, 2021), Johnson & Johnson (March 12, 2021), and Sinopharm (May 7, 2021) vaccines.

The development and marketing of vaccines against SARS-CoV-2 are for WHO and governments one effective solution to limit the ability of the pathogen to spread **[4]**. Indeed, thanks to the so-called “herd”, “indirect”, or “group” immunitý **[4]**, vaccination allows individuals and communities to remain protected and to decrease the probability of an outbreak. For example, the governments of Benin and Senegal launched their COVID-19 vaccination campaigns on February 23, 2021, and March 29, 2021, respectively. However, as of December 4, 2022, only three countries in Africa have reached the target of 70% of their population fully vaccinated according to the WHO Strategy **[5]**: Seychelles (76.7%), Liberia (79.9%) and Mauritius (86.0%). By December 2022, 20.9% of the population is fully vaccinated in Benin, while in Senegal (7.6%), the 10% threshold has not yet been exceeded **[5]**. This low vaccination coverage observed in both countries does not eliminate the possibility of epidemic transmission with the presence of a reservoir of unvaccinated individuals susceptible to infection. What are the motivations of Beninese and Senegalese to accept being vaccinated? Recent studies have shown that concerns about perceived safety and efficacy **[6, 7, 8]**. or lack of reliable information about vaccines **[9, 10]** were the main barriers to adoption of SARS-CoV-2 vaccines. However, these works use classical techniques with limitations for modeling vaccine intention or hesitancy, a complex decision-making process with multiple sources of influence **[11, 12]**. In fact, in contrast to these classical methods, structural equation models can fulfil the following conditions **[13]**: ability to handle simultaneously several sets of observed explanatory and explained variables (thus the stage of causal relationships), ability to analyze the links between the different dimensions and to consider errors at the level of measurement (reduction of psychometric biases), and finally, the ability of confirmatory applications.

Structural equation modeling was adopted to better understand and interpret decision-making on vaccination against COVID-19 in Benin and Senegal.

## SCOPE OF THE STUDY

Senegal is in West Africa with 14 administrative regions. The population of Senegal in 2019 is estimated at 16,209,125 and is predominantly female (50.2%). The ratio of telephone numbers per person is 1.1. The proportion of people using a cell phone at least five times a day increased from 36.42% in 2014 to 73.46% in 2017 **[14]**. With a population of 11,496,140, 50.9% of whom are women, Benin is a West African country. In 2021, cell phone penetration was estimated at 101.8%, which corresponds to a mobile subscriber base of 12,731,782 **[15]**.

## METHODS

### Type of Study

This was a cross-sectional, descriptive, and analytical nation-wide study remotely conducted via telephone from a call center. Data were collected from December 24, 2020, to January 16, 2021, for Senegal and from March 29 to May 14, 2021, for Benin.

### Study population

The study population consisted of the population of Senegal and Benin, aged 18 years and above, proportionally distributed according to age, sex, and region.

### Sampling

A marginal quota survey was conducted **[16]**. This method is relevant in emergencies such as the COVID-19 pandemic with sample sizes below 3000 **[16,17]**. An appropriate choice of quotas can reduce the estimate’s variance and the magnitude of its confidence interval. If done rigorously, the quota sampling method can be as accurate as random sampling **[17]** or better if the sample size is small **[18,19]**. The following variables were used to define the quotas: age, gender, and region **[20]**. In June 2020, we conducted a first nationwide telephone survey of 813 people in Senegal to measure the social acceptability of governmental measures to control COVID-19 **[21]**. Based on this first survey, which did not concern the vaccine aspects, we organized a second survey among these people. The final sample size in Senegal was 607 (74.6%). A comparison of the characteristics of the quotas chosen to constitute the sample between the two surveys shows that they are not statistically different for region (p = 0.99) and age (p = 0.08) but are for gender (p = 0.04). In Benin, this study was a first and a total of 865 people could answer all the necessary questions.

### Data collection

A system based on the Random Digit Dialing (RDD) method **[22]** was set up to collect the data. The first step consisted in generating random numbers according to the market shares of the different operators present in each country. A computer program was used to identify valid numbers by sending mass SMS messages based on the delivery status of the SMS messages. This system, networked with an automated call center, automatically dialed the valid numbers and put them in touch with a human agent at the call center who explained the research and asked for consent to participate. From that moment on, the call center agent proceeded to fill in the digitalized questionnaire with the Open Data Kit (ODK) software and integrated it into the collection system.

After three days of training on the protocol and content of the survey, data collection was carried out by interviewers who spoke, in addition to French, the five leading national languages in Benin (Fon, Yoruba, Bariba, Dendi, and Adja) and Senegal (Wolof, Pulaar, Serer, Mandingo, and Diola). Data quality assurance (DQA) covered all phases of the study: before data collection began, during data collection, and after data collection (Table 2).

**Table 2:**
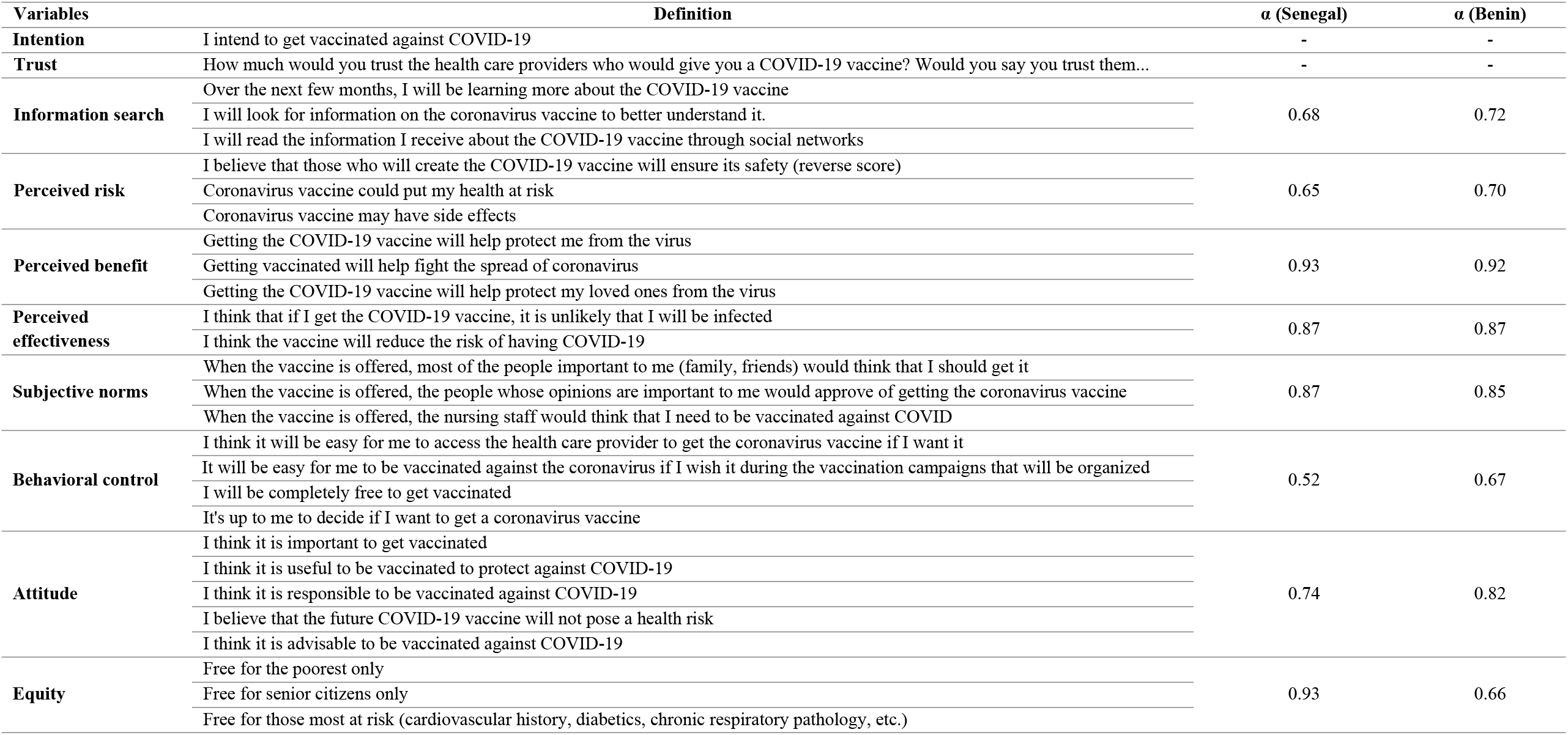
Dimensional items and internal consistency

**Table 2:** Data Quality Assurance Procedure

|  | **Before the**<br>**start of data**<br>**collection** | **During data**<br>**collection** | **At the end**<br>**of the data**<br>**collection** |
| --- | --- | --- | --- |
| National Health Research Ethics Committee approval, standardization of collection procedures during interviewer training and pre-testing of tools | ✓ |  |  |
| Supervision of collection, verification of compliance with collection protocol and procedures, reviews of 10% of completed forms per enumerator |  | ✓ |  |
| Consistency check, checking for missing values and outliers |  | ✓ | ✓ |

Prior to data sharing, all data is de-identified and separated from identifying information such as geographic location, names, cell phone numbers or other identifying information of respondents. It is only the domain identification codes that are included in the electronic data files, to maximize the preservation of respondent confidentiality.

### Theoretical and conceptual frameworks

The design of the study questionnaire is based on two theoretical models chosen for their relevance to the research question: the Theory of Planned Behavior (TPB) **[23]** and the Health Belief Model (HBM) **[24,25]**. For the former, health behavior is related to attitudes, subjective norms and perceived behavioral control. As for the second model, health behavior would be explained by the perception of the seriousness of the disease, the effectiveness, and the perceived benefits of preventive action, but also the perceived risks. For each dimension, we tested the reliability of the measurement instruments using Cronbach’s alpha. They were all above 0.6 (Table 1), and, therefore reliable **[26]**.

✓ **Intention to be vaccinated**. This variable was measured by a question (“I intend to be vaccinated against COVID-19”). Responses were defined according to a 5-point Likert scale (“Strongly agree=5” to “Strongly disagree=1”)
✓ **Trust** It was measured by the question (“How much would you trust healthcare providers who would give you a COVID-19 vaccine?
✓ **Information search** It was measured by three items related to information seeking about the vaccine (e.g., “Over the next few months, I will regularly learn about the COVID-19 vaccine”).
✓ **Perceived risk** It was measured by 3 items (e.g., “I believe that those who are going to create the COVID-19 vaccine will ensure its safety”)
✓ **Perceived benefit** This dimension consisted of 3 questions (e.g., “Getting the COVID-19 vaccine will help protect me from the virus”)
✓ **Perceived effectiveness** It was measured by two items (e.g., “I think that if I get vaccinated against COVID-19, I am unlikely to be infected”).
✓ **Subjective norms** This scale was measured by 3 items (e.g., “When the vaccine is offered, most people important to me (family, friends) would think I should get the COVID-19 vaccine”)
✓ **Behavioural control** This scale was measured by 4 items (e.g., “I think it will be easy for me to access the health care provider to get the coronavirus vaccine if I want it”)
✓ **Attitude** This scale was measured by five items (e.g., “I think it is important to get vaccinated”)
✓ **Equity** This scale was measured by 4 items (e.g., “Free for seniors only”)

Each item was measured on a 5-point Likert scale (1 = “Strongly Disagree”, 5 = “Strongly Agree”). The first item of the perceived risk dimension was scored inversely.

### Data analysis

The data analysis is performed in three steps.

A descriptive analysis of the variables under study (dependent and independent). Qualitative data were described through frequency measures with confidence intervals with a 95% confidence level and quantitative data through central tendency and dispersion measures.

A cross-tabulation analysis between the dependent variable and the relevant independent variables. Appropriate statistical tests (Student, Mann-Whitney) were used to measure associations at 5% alpha risk. Correlations were also performed to investigate the relationships between the different dimensions **[27]**.

A confirmatory factor analysis based on structural equation modeling tested the reliability, validated the scales (convergence and discrimination) and also identified factors associated with the intention to be vaccinated against COVID-19. This analysis estimated complex relationship models (multiple independent and dependent variables) that eventually incorporated latent variables (constructed from multi-item measurement scales) **[28]**. The different measures have good goodness of fit: the Comparative Fit Index (CFI) > 0.90; the Root Mean Square Error of Approximation (RMSEA) < 0.10 and the Standardized Root Mean Residual (SRMR) < 0.08 **[29,30]**. The internal consistency reliability of the model was assessed through Cronbach’s alpha, with a desired level of > 0.60 **[26,31]**.

All analyses were performed with R software **[32]**.

### Ethical considerations

The research received approval from the National Health Research Ethics Committee of Senegal (SEN/20/23) and the Local Ethics Committee for Biomedical Research of the University of Parakou in Benin (0308/CLERB-UP/P/SP/R/SA). All individuals were informed of the ethical issues and the possibility of withdrawing from the study at any time. They all consented to participate.

## RESULTS

The age ranges from 25 to 59 years, was the majority in Senegal (67.1%), while the age range under 25 years was the majority in Benin (57.6%). Men predominated with 60.3% in Senegal and 59.2% in Benin (Table 3).

**Table 3:** Distribution of individuals by socio-demographic characteristics and intention to vaccinate.

| Characteristics | Senegal |  |  |  | Benin |  |  |  |
| --- | --- | --- | --- | --- | --- | --- | --- | --- |
|  | Headcount | Proportion |  | [95% CI] | Headcount | Proportion |  | [95% CI] |
| Male | 366 | 54.1% | 49.0% | 59.2% | 350 | 68.4% | 64.2% | 72.3% |
| Female | 241 | 54.8% | 48.4% | 61.0% | 210 | 59.5% | 54.3% | 64.5% |
| < 25 years | 140 | 50.0% | 41.8% | 58.2% | 324 | 65.1% | 60.8% | 69.1% |
| 25-59 years | 407 | 54.5% | 49.7% | 59.3% | 207 | 64.9% | 59.5% | 69.9% |
| >=60 years | 60 | 63.3% | 50.5% | 74.5% | 29 | 60.4% | 46.1% | 73.2% |
| No education | 253 | 57.3% | 51.1% | 63.3% | 98 | 70.5% | 62.4% | 77.5% |
| Primary | 122 | 56.6% | 47.6% | 65.1% | 128 | 71.1% | 64.1% | 77.3% |
| Secondary | 153 | 51.6% | 43.7% | 59.5% | 229 | 65.8% | 60.6% | 70.6% |
| Tertiary | 79 | 46.8% | 36.1% | 57.8% | 105 | 53.0% | 46.1% | 59.9% |

In Senegal, vaccine intention was estimated at 54.4% and was higher among the elderly (63.3%) and the uneducated (57.3%). In Benin, the intention to vaccinate was 64.7% and was higher among the less educated (70.3%) and young people under 25 (65.1%).

The internal consistency reliability of the TPB constructs was good, all above the 0.60 threshold **[24]**, except for the behavioral control dimension in Senegal (α=0.52) (Table 2). Significant positive correlations existed between the dimensions, with Pearson correlation coefficients ranging from 0.14 (Equity and Perceived Efficacy) to 0.68 (Attitude and Perceived Benefit). Correlations between all constructs (except for perceived risk) and intention were also positive, as expected, with correlations ranging from relatively low (r=0.08 and r=0.03; Equity and Intention (Benin and Senegal)) to high (r=0.67 and r=0.55; Attitude and Intention (Benin and Senegal)) (Table 4).

**Table 4:** Correlation matrix

| SENEGAL |  |  |  |  |  |  |  |  |  |  |
| --- | --- | --- | --- | --- | --- | --- | --- | --- | --- | --- |
|  | 1 | 2 | 3 | 4 | 5 | 6 | 7 | 8 | 9 | 10 |
| Intention (1) | - |  |  |  |  |  |  |  |  |  |
| Trust (2) | 0.52*** | - |  |  |  |  |  |  |  |  |
| Information search (3) | 0.20*** | 0.17*** | - |  |  |  |  |  |  |  |
| Perceived risk (4) | -0.59*** | -0.40*** | -0.10 | - |  |  |  |  |  |  |
| Perceived benefit (5) | 0.74*** | 0.57*** | 0.17*** | -0.61*** | - |  |  |  |  |  |
| Perceived effectiveness (6) | 0.65*** | 0.56*** | 0.16*** | -0.55*** | 0.76*** | - |  |  |  |  |
| Subjective standards (7) | 0.66*** | 0.56*** | 0.20*** | -0.51*** | 0.67*** | 0.65*** | - |  |  |  |
| Behavioral control (8) | 0.32*** | 0.39*** | 0.25*** | -0.24*** | 0.38*** | 0.34*** | 0.32*** | - |  |  |
| Attitude (9) | 0.67*** | 0.44*** | 0.27*** | -0.52*** | 0.68*** | 0.61*** | 0.58*** | 0.27*** | - |  |
| Equity (10) | 0.08 | 0.24*** | 0.06 | -0.08 | 0.16*** | 0.14*** | 0.15** | 0.16*** | 0.07 | - |
| BENIN |  |  |  |  |  |  |  |  |  |  |
|  | 1 | 2 | 3 | 4 | 5 | 6 | 7 | 8 | 9 | 10 |
| Intention (1) | - |  |  |  |  |  |  |  |  |  |
| Trust (2) | 0.42*** | - |  |  |  |  |  |  |  |  |
| Information search (3) | 0.18*** | 0.28*** | - |  |  |  |  |  |  |  |
| Perceived risk (4) | -0.51*** | -0.36*** | -0.10* | - |  |  |  |  |  |  |
| Perceived benefit (5) | 0.55*** | 0.64*** | 0.33*** | -0.42*** | - |  |  |  |  |  |
| Perceived effectiveness (6) | 0.39*** | 0.88*** | 0.28*** | -0.29*** | 0.63*** | - |  |  |  |  |
| Subjective standards (7) | 0.59*** | 0.45*** | 0.28*** | -0.40*** | 0.60*** | 0.43*** | - |  |  |  |
| Behavioral control (8) | 0.37*** | 0.47*** | 0.44*** | -0.23*** | 0.51*** | 0.48*** | 0.48*** | - |  |  |
| Attitude (9) | 0.55*** | 0.53*** | 0.31*** | -0.44*** | 0.66*** | 0.49*** | 0.52*** | 0.49*** | - |  |
| Equity (10) | 0.03 | -0.04 | 0.08 | -0.09 | 0.04 | -0.09 | -0.03 | 0.01 | 0.02 | - |

\*\*\* p < 0.001; \*\* p < 0.01 ; \* p < 0.05

**Table 5:**
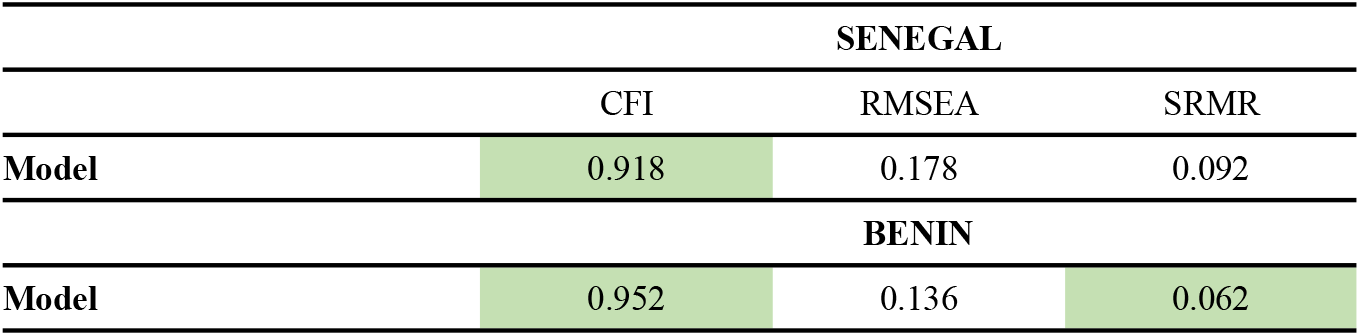
Structural equation model fit index

Results show that in Senegal, intention to vaccinate is positively associated with a good perception of the benefits of vaccination (β=0.33***), a positive attitude (β=0.22***) towards vaccines, and sensitivity to subjective norms (β=0.19***) (Figure 1). Low trust in health care providers (β= -0.40***) amplified the influence of perceived risk (β= -0.14***) on the intention not to vaccinate (Figure 1). In other words, when trust in healthcare providers is low, the influence of perceived disease risk on the decision to reject vaccination is greater. Aspects related to behavioral control, information seeking, efficacy, or equity do not appear to be correlates of vaccine intention. However, perceived vaccine efficacy was affected by perceived disease risk (β=-0.12***) and improved with reasonable apprehension of the benefits of vaccination (β=0.60***) (Figure 1).

**Figure 1:**
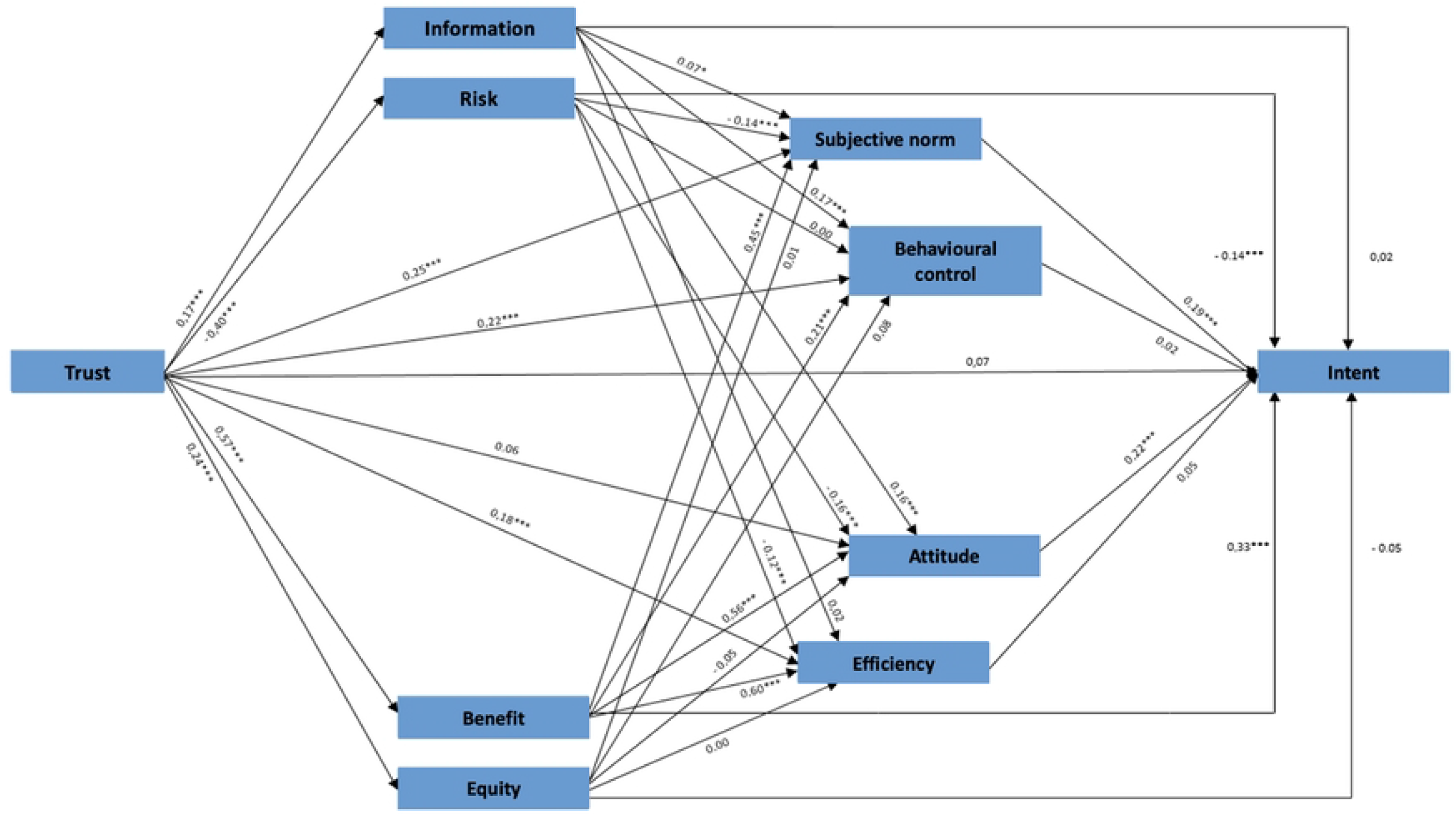
Structural equation of the intention to vaccinate model for COVID-19 (Senegal)

In Benin, the same trends are observed as intention to vaccinate is positively associated with attitudes (β=0.20***), subjective norms (β=0.32***) and perceived benefits (β=0.12***) which is negatively associated with perceived risks (β= -0.25***) (Figure 2).

**Figure 2:**
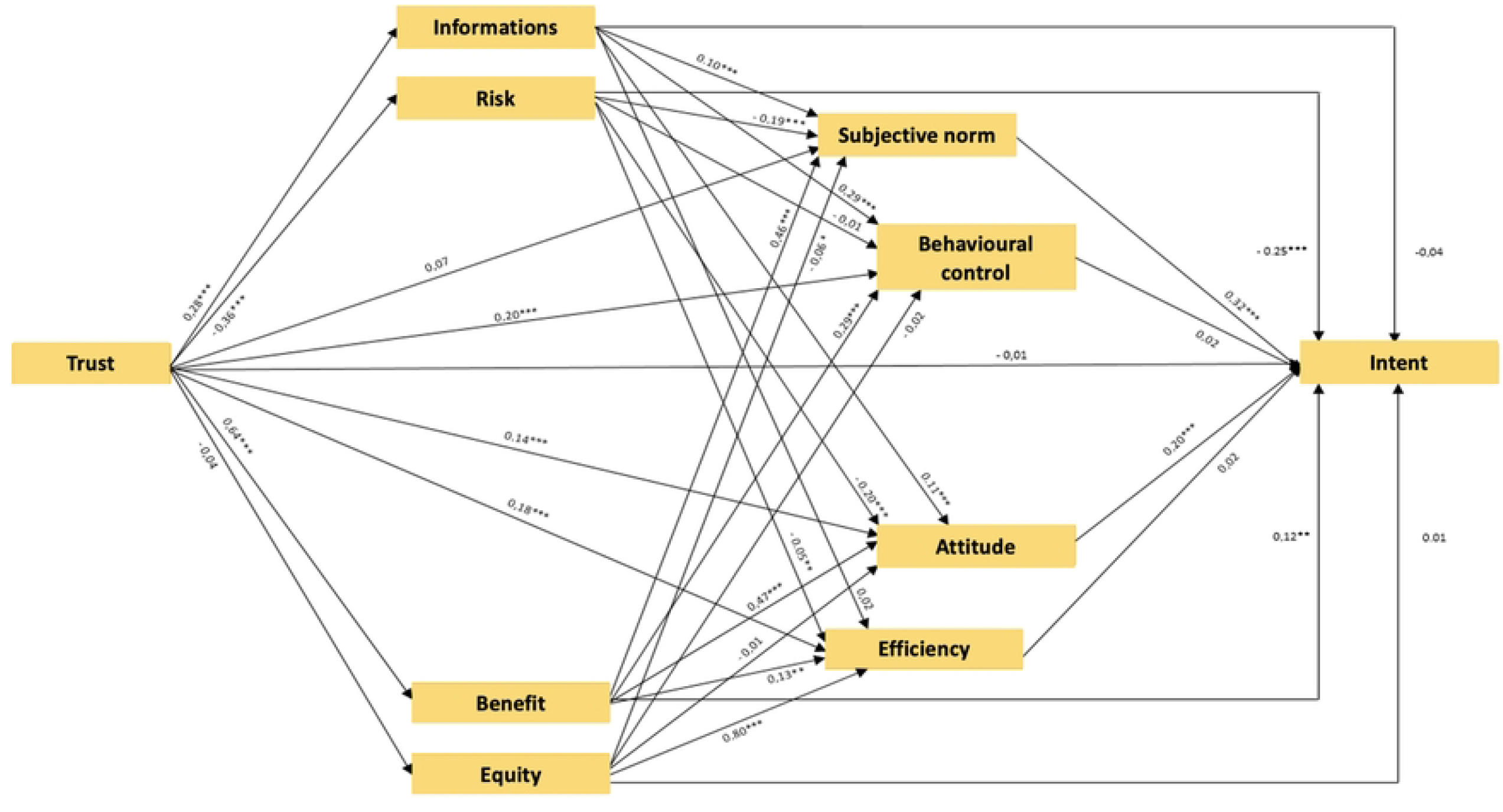
Structural equation of the intention to vaccinate model for COVID19 (Benin)

## DISCUSSION

The results show that the **perceived risk** of vaccines negatively influences the intention to be vaccinated. The origin of the rejection or hesitation lies in a higher perceived risk of side effects, low confidence in vaccines (safety) or to avoid a hypothetical and delayed risk of disease. Numerous studies indicate that the lack of perception of long-term side effects **[33]**, the accelerated development of vaccines and the wave of false or misleading information on social networks and search engines **[35,36,37]** affect confidence in these vaccines and, therefore adherence to vaccination. Risk perception is built through exposure to false information sometimes conveyed in social networks, which are widely used in Africa **[38]**. The latter, using algorithms, favor attractive content, even if it is repeatedly denied by the scientific community. Indeed, distrust of COVID-19 vaccines is the result of misinformation and fake news, mainly on social media **[38]**. To address misinformation and facilitate the acceptability of COVID-19 vaccines, strategies to provide accurate information by emphasizing the reasons and importance of vaccination should be implemented. Such a strategy has been initiated in the Netherlands, where a think tank on misinformation has been set up. Experts debunk misinformation about the vaccine on a voluntary basis using their own personal social media accounts **[39]**.

Regarding **attitude**, results show that a positive perception of vaccination significantly impacts the intention to vaccinate. In other words, people who are favourable to vaccination are generally those who think that it is important, valuable, responsible, or desirable to be vaccinated. These are also the people who believe that vaccines do not pose a health risk. These results are like those obtained by Greyling and Rossouw **[40]** who, using a Vaccine Positive Attitude Index (VPAI) created from vaccine-related tweets collected in real time in 10 countries (Australia, Belgium, Germany, Great Britain, France, Italy, the Netherlands, New Zealand, South Africa, and Spain) between February 1 and July 31, 2021, showed that positive attitudes are associated with vaccine adoption. Their findings also showed that information about safety and expected side effects are likely to improve positive attitudes toward COVID-19 vaccines **[40]**. Indeed, a study conducted in Senegal also showed that a poor attitude towards vaccination was significantly related to vaccine hesitancy [6]. In addition, a systematic review and meta-analysis examining Health Community Workers’ (HCWs) willingness to accept COVID-19 vaccination identified, among other things, a positive attitude towards the vaccine as a factor associated with increased desire to be vaccinated **[41]**.

Regarding the **perceived benefit**, the results show that a positive perception of the benefits of vaccination is a correlate of the intention to be vaccinated. The perception that vaccination provides individual and collective protection is a factor of adherence. These results confirm two components of the 5C model **[42]** that was used in 2021 by the European Centre for Disease Prevention and Control (ECDC) to diagnose the causes of poor vaccine uptake and low vaccination coverage by analyzing cross-sectional data on the Finnish population **[43]**. The first component shows that people who perceive a vaccine-preventable risk of infection are those with a solid desire to vaccinate **[44-45]**. Also, complacency affects individual perception of vaccine efficacy even if it is not a direct determinant of vaccine adherence. The second component is collective responsibility, which refers to the willingness of individuals to participate in the fight against the spread of the virus in a collective effort to achieve mass immunity **[45]**. Vaccination thus becomes a civic act, essential to obtain sufficient vaccine coverage and joint protection to stop the disease.

The results also show that **subjective norms**, reflected in the approval of the vaccine by significant others, affect the intention to vaccinate. In other words, vaccination decisions would be influenced by the positions of significant others (family, friends, health providers, etc.). Three groups of researchers using the Theory of Planned Behavior (TPB) conceptual framework have reached similar conclusions **[46,47,48]**. In addition, a qualitative analysis of the perception of vaccination by the vaccinated elderly subject showed that the sources of motivation to be vaccinated were the family via affect and the treating physician as a scientific referent **[49]**.

Based on these findings, it would be necessary for the Senegalese and Beninese authorities in charge of public health to implement information campaigns aimed at encouraging positive attitudes toward the vaccine with key messages translated into several languages. Therefore, they should encourage individuals to share positive attitudes and experiences about vaccination with those around them. Also, because individuals do not necessarily have the medical skills to make decisions **[48]**, health professionals are key stakeholders in improving immunization coverage. Therefore, communicating on the social benefits of vaccination is an efficient way to increase vaccination intention. Indeed, the perception of serving their community and loved ones by getting vaccinated is a unifying factor for the population around vaccination. In addition, younger people’s adherence to vaccination may be greater if they perceive it as protection for their elderly family members. Such a strategy could be based on key messages such as “vaccination to protect oneself while protecting others” or “vaccination is the solution for a return to normal life” **[50]**.

## LIMITS

The samples were only nationally representative and did not allow for disaggregation by residence or region. Only people with a cell phone were interviewed, thus excluding the most marginalized populations.

## CONCLUSION

Understanding people’s expectations, concerns, and beliefs about COVID-19 and the vaccine is fundamental to the success of vaccination campaigns. In both Benin and Senegal, the results showed that a good perception of the benefits of vaccination, a positive attitude, and sensitivity to subjective norms positively influenced the intention to be vaccinated. Also, low trust in healthcare providers amplified the perceived risk of vaccination, which negatively impacted the intention to vaccinate. These results will certainly better inform vaccination strategies, particularly by considering the causal relationships between the different dimensions. However, in-depth studies will be necessary to understand and identify the enclaves of low vaccine coverage in both countries (<25%) despite a relatively high intention to vaccinate (>50%). Could these low rates be explained by the fact that campaigns do not consider causal relationships between these different dimensions? But also, perhaps because it is not useful to vaccinate, and people know it? Many questions that these in-depth analyses can explore…

## Data Availability

All data are included in the paper.

## FINANCING

This research is part of the Support to the African Response to the COVID-19 Epidemic (ARIACOV) program funded by the French Development Agency (AFD).

## ANNEXES

